# Multivariate genetic analysis maps shared biology between psoriasis and cardiovascular diseases

**DOI:** 10.64898/2026.10.19.26365878

**Authors:** Charalabos Antonatos, Eleftheria Georgalaki, Ioanna Tzoulaki, Yiannis Vasilopoulos

## Abstract

Psoriasis is associated with increased cardiovascular morbidity, but the extent and structure of the shared genetic architecture remain incompletely understood. We integrated large-scale European-ancestry genome-wide summary statistics for psoriasis and seven cardiovascular diseases to quantify genetic overlap, model shared liability and prioritize candidate mechanisms. Psoriasis showed modest genetic correlations with all cardiovascular outcomes, but mixture modelling estimated that more than half of psoriasis-associated non-zero variants were shared with five cardiovascular diseases. Genomic structural equation modelling of a latent psoriasis-CVD factor identified 167 independent risk loci, including 50 not genome-wide significant in any component trait. Multimodal prioritization nominated 473 candidate genes and 90 approved compounds with therapeutic relevance, including shared targets involving *PDE4B*, *GLP1R*, *KEAP1* and *PRDM16*. Mediation and Mendelian randomization analyses implicated cardiometabolic traits, although no single trait accounted for more than half of the shared liability. These results prioritize genetic mechanisms and pharmacological hypotheses for psoriasis-associated cardiovascular multimorbidity.

## Introduction

Psoriasis is a chronic inflammatory skin disease affecting approximately 2-3% of the global population [1]. Genetic studies indicate that psoriasis has a polygenic architecture with systemic consequences beyond the skin [2]. Cardiovascular disease (CVD) is one of its most important comorbidities. Pooled analyses of more than 600000 patients have reported 15-45% higher relative risks of cardiovascular outcomes, with 40-70% excess risk in severe disease requiring systemic therapy [3,4]. This risk persists after adjustment for conventional cardiometabolic risk factors [5], indicating that shared risk factors explain only part of the psoriasis-CVD relationship. One proposed explanation is the psoriatic march, in which chronic inflammation links skin disease to cardiometabolic and vascular pathology [6]. Coronary imaging studies further support similar elements, where patients with psoriasis show increased non-calcified and high-risk coronary plaque burden [7]. In some cohorts, administration of modern biologics targeting central cytokines in psoriasis reduces circulating inflammatory biomarkers and vascular inflammation [8]. Given the availability of effective treatments targeting inflammatory pathways in psoriasis, determining the extent to which psoriasis and CVD share underlying biological mechanisms may help identify pathways and candidate targets relevant to both conditions [9].

Genome-wide association studies (GWASs) have further demonstrated that both psoriasis [2] and CVDs [10] are highly polygenic, with psoriasis risk enriched immune-related signals and CVD susceptibility involving vascular, metabolic and inflammatory pathways. Linkage disequilibrium score regression (LDSC) and trans-disease meta-analysis of psoriasis and coronary artery disease have uncovered evidence for shared heritable signals [11], while Mendelian randomization evidence suggests that genetic liability to CVDs may increase psoriasis risk [12]. More recent studies extending across multiple cardiovascular outcomes have reported pleiotropic loci and candidate genes enriched in immune and vascular tissues and converging on T-cell-mediated immune dysregulation [13]. A deeper understanding of this shared genetic architecture could provide insights into common etiological mechanisms and prioritize candidates for therapeutic investigation.

Here, we examined the genetic basis of psoriasis-associated cardiovascular multimorbidity using large-scale European-ancestry GWAS summary statistics for psoriasis and seven major CVDs [3,14,15]. First, we combined genome-wide and local correlation analyses with mixture modelling to characterize the extent and structure of polygenic overlap. Second, we performed a multivariate GWAS through genomic structural equation modeling (SEM) [16] and a series of post-GWAS analyses to identify possible genes, tissues and biological mechanisms that may underlie this shared liability. Finally, we used mediation analyses and bidirectional Mendelian Randomization (MR) to evaluate which modifiable risk factors potentially mediate the psoriasis-CVD multimorbidity.

## Results

Our approach is presented at Fig. 1. We gathered summary statistics from the largest GWASs in Europeans in psoriasis [2] and CVDs with epidemiological evidence for association with psoriasis, including atrial fibrillation (AF) [17], coronary artery disease (CAD) [10], heart failure (HF) [2], hypertension (HT) [18], myocardial infarction (MI) [19], peripheral artery disease (PAD) [20] and stroke [21]. Details for all GWASs are presented at Table S1.

**Fig. 1.**
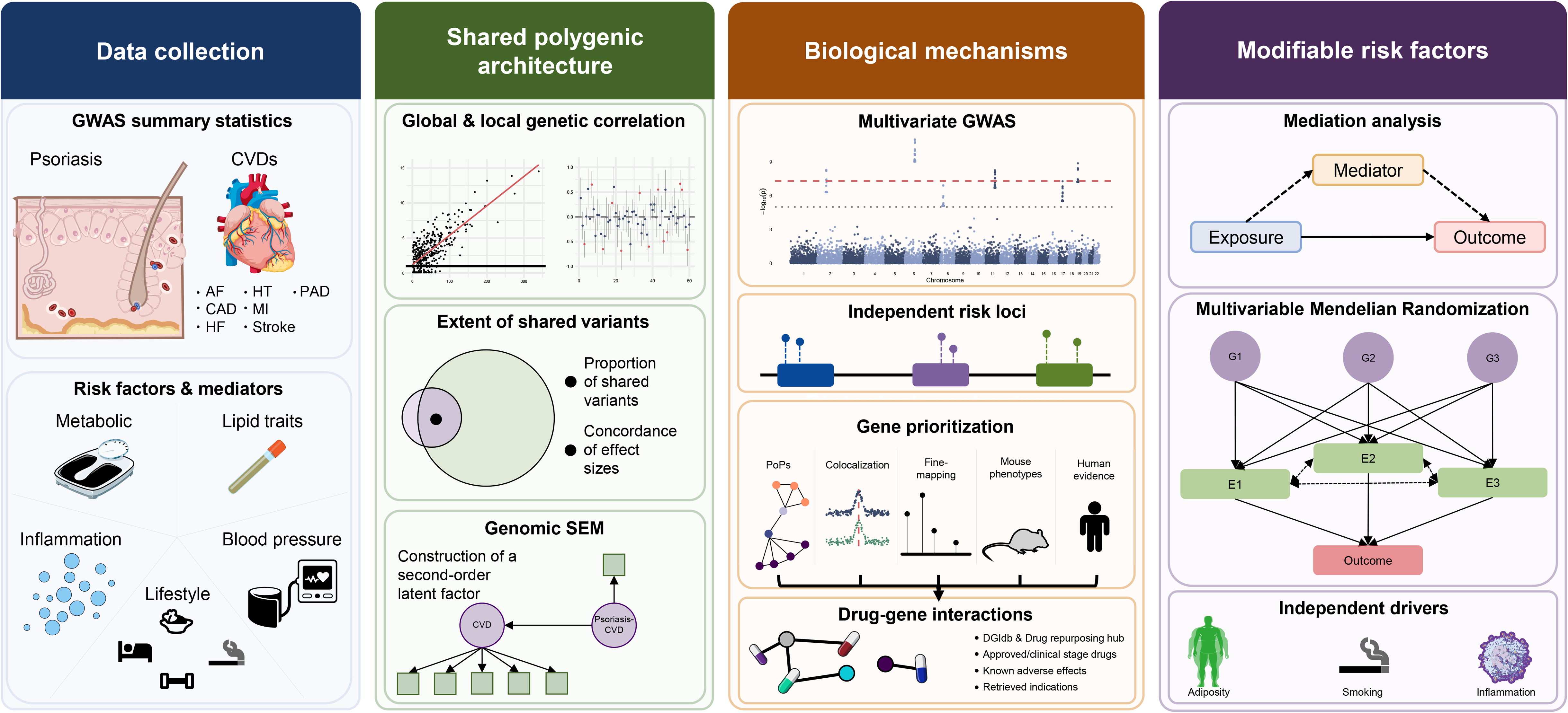
Study design.

### Extent and structure of shared genetic architecture

At the genome-wide level, psoriasis showed modest but significant genetic correlations with all CVDs examined (Fig. 2a), ranging from 0.078 (0.023-0.133) with AF to 0.256 (0.173-0.3) with PAD (Table S2). Regional-level analysis identified 29 significant psoriasis-CVD local genetic correlations across 2,495 approximately independent genomic regions (Table S3) [36]. The strongest regional overlap mapped to the pleiotropic SH2B3-ATXN2 locus at 12q24 was positively correlated between psoriasis and 3 CVDs. At the major histocompatibility complex (MHC), correlation estimates were mildly negative for 2 CVDs (Fig. 2b).

**Fig. 2.**
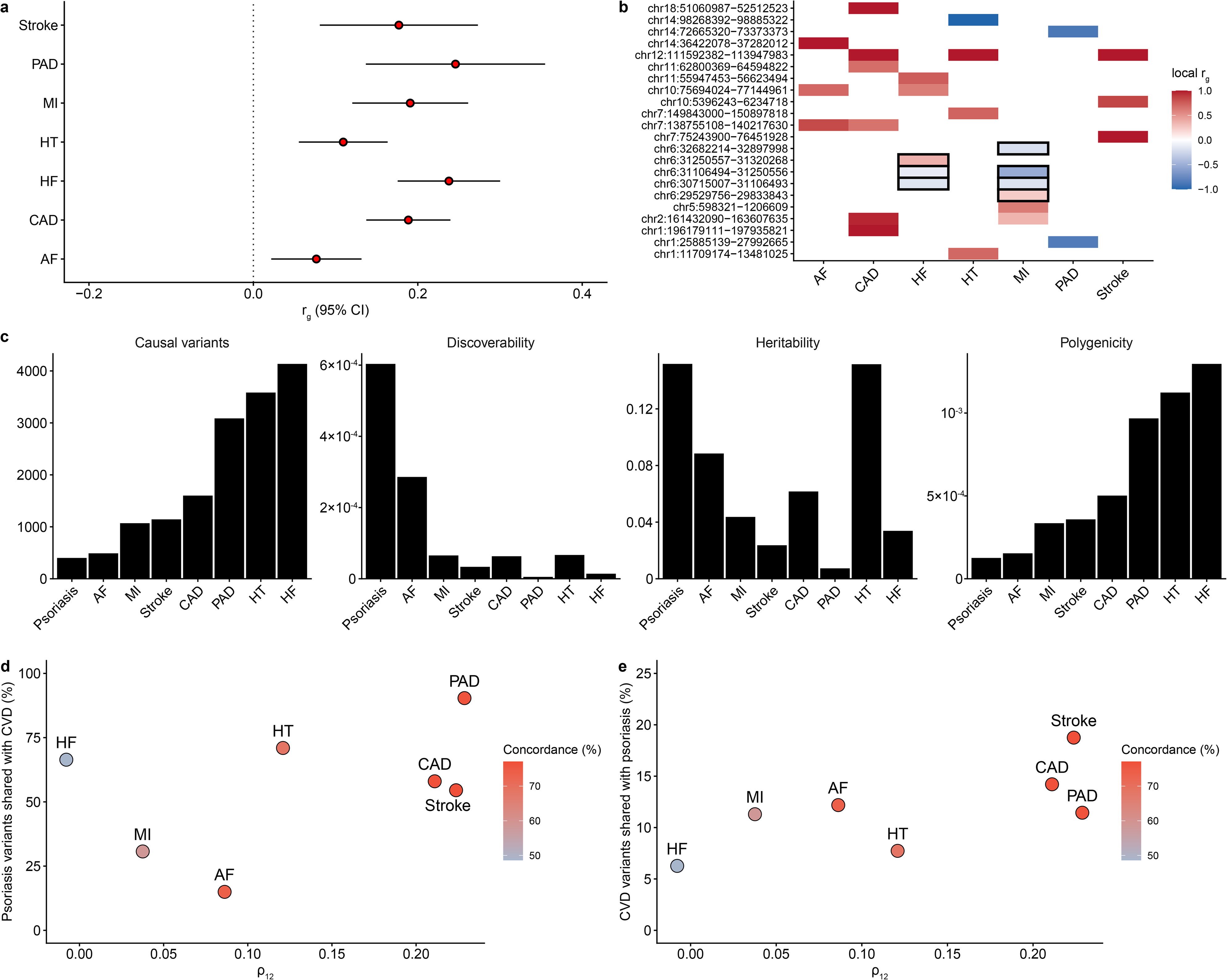
Shared genetic architecture between psoriasis and cardiovascular disease traits. (a) Genome-wide genetic correlations (r_g_) between psoriasis and cardiovascular disease (CVD) traits estimated using LD Score Regression. Points show r_g_ estimates with 95% confidence intervals. Red fill indicates associations significant after Bonferroni correction (P-value<0.05/7). (b) Local genetic correlations between psoriasis and CVD traits at genomic loci showing significant local r_g_ using LAVA. Tile color represents the local genetic correlation coefficient with red indicating positive and blue indicating negative correlation. Loci are ordered by chromosomal position and labeled by genomic coordinates. Loci within the MHC region (chromosome 6) are outlined in black. Only loci reaching locus-wise FDR-corrected significance are shown. (c) Univariate MiXeR estimates of polygenic architecture for psoriasis and each CVD trait showing discoverability (σ²β), polygenicity (π), estimated number of causal variants and SNP-based heritability (h²). Bars are ordered by estimated value within each panel. (d, e) Bivariate MiXeR analysis of polygenic overlap between psoriasis and each CVD trait. (d) Percentage of psoriasis non-zero variants shared with each CVD trait plotted against the genome-wide genetic correlation estimated by MiXeR. (e) Percentage of CVD non-zero variants shared with psoriasis plotted against genetic correlation estimated by MiXeR. In both panels, point fill indicates the percentage of concordant effects among shared non-zero variants.

Although genetic correlation estimates were modest, they quantify direction and magnitude of genetic effects rather than the extent of overlap. Hence, we applied MiXeR to investigate univariate polygenicity (Table S4) and bivariate pleiotropic architecture (Table S5) [37]. Psoriasis displayed the lowest polygenicity with the fewest variants exhibiting non-zero effects (n=389±32) and the smallest number of variants explaining 90% of the trait’s heritability (h^2^). In contrast, psoriasis demonstrated the highest discoverability, defined as the variance of effect sizes among non-zero variants, and the largest single-nucleotide polymorphism (SNP)-based h^2^ estimate (Fig. 2c). Despite this sparse architecture, bivariate MiXeR revealed extensive sharing between psoriasis and CVDs. More than half of psoriasis-associated non-zero variants were shared with 5/7 CVDs with large concordance estimates (Fig. 2d), spanning from 54.52% shared with stroke (77.06% concordant) to 90.38% shared with PAD (76.53% concordant). The shared polygenicity pattern was markedly asymmetric when comparing non-zero variants of CVDs, among which the fraction shared with psoriasis ranged from 18.75% for stroke to 6.26% for HF (Fig. 2e). Performance metrics supported the robustness of all inferred polygenic overlaps validating that the estimates exceeded the minimum expected level while remaining distinct from the maximum possible overlap scenario (Table S5). Overall, these results highlight that psoriasis-associated variants are highly enriched for variants contributing to cardiovascular traits.

### Functional characterization of multimorbidity

Having established the extensive but asymmetric genetic overlap, we sought to identify the genomic basis of multimorbidity within a common framework using genomic SEM [16]. We first formed a CVD latent factor including CAD, HF, HT, PAD and stroke, excluding AF and MI which deteriorated the model fit. The loading of HF in the latent CVD factor was set to 1, reporting a comparative fit index (CFI) of 0.988 and SRMR of 0.044. We then fitted psoriasis as an indicator of a second-order latent psoriasis-CVD factor (Fig. 3a). This model showed an acceptable model fit (CFI=0.986, SRMR=0.0406), implying that a single latent factor provides an acceptable representation of the genetic covariance between psoriasis and included CVDs. The loading of psoriasis in the latent psoriasis-CVD factor was fixed to 1, and the latent CVD factor loaded with 3.436 [2.612-4.261 (P-value=3.03×10^-16^)] in the multimorbidity factor.

**Fig. 3.**
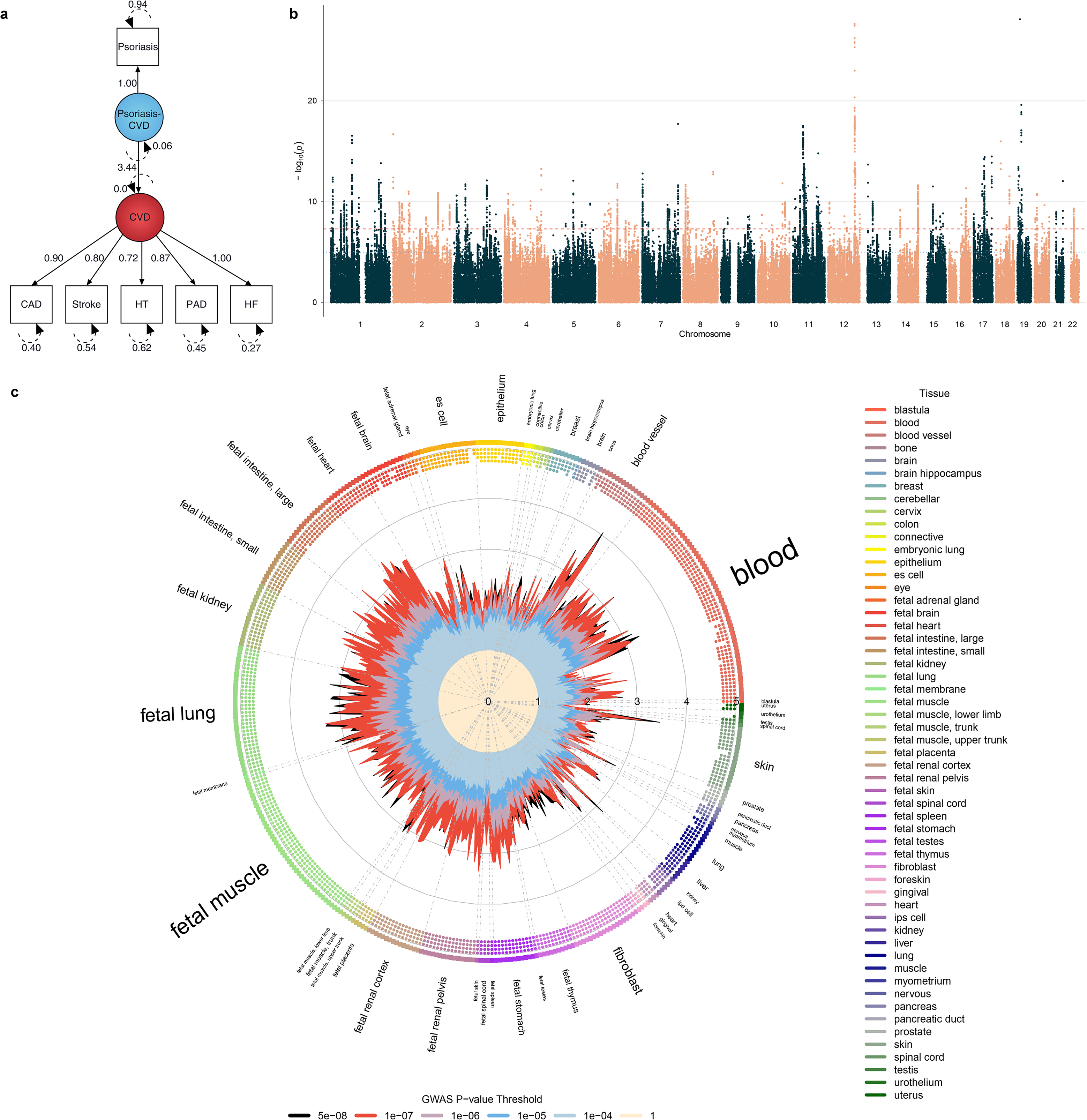
Genomic structural equation modeling of a shared psoriasis-CVD factor and genome-wide association signals. (a) Path diagram of the common-factor model fit using Genomic SEM. A latent cardiovascular disease (CVD) factor was defined by heart failure, coronary artery disease, stroke, hypertension, and peripheral artery disease and a second-order latent factor (psoriasis-CVD) was defined by psoriasis and the CVD factor. Edges show standardized loadings. The observed variables are shown in rectangles and the latent variables in circles. (b) Manhattan plot of the genome-wide association study for the shared psoriasis-CVD latent factor. (c) GARFIELD functional enrichment analysis of psoriasis-CVD-associated variants within DNase I hypersensitivity sites, showing enrichment of the shared genetic signal in regulatory/open-chromatin regions. Enrichment was conducted for P-value≤5×10^-8^, 10^-7^, 10^-6^, 10^-5^, 10^-4^ and 1 thresholds.

We performed a multivariate GWAS on the latent psoriasis-CVD factor across 3564815 variants. Approximately 10.3% of variants showed evidence of SNP-specific heterogeneity from the common-factor model (Q_SNP_ <0.05). Excluding variants with significant Q_SNP_ heterogeneity resulted in 167 independent genomic risk loci (Fig. 3b). The LDSC attenuation ratio was 0.107, suggesting that inflation in association statistics (λ_GC_=1.72) was largely driven by polygenicity. Among these loci, 50 were not genome-wide significant in any constituent GWAS, suggesting that multivariate modelling can identify associations not detected in individual traits (Table S6). The latent psoriasis-CVD factor showed expected correlation patterns with all involved CVDs (Table S7) and psoriasis was positively associated with the latent factor (0.518, 0.44-0.595). Across P-value thresholds, variants associated with the latent factor were enriched in regulatory regions across multiple tissues and biological contexts (Table S8) [38]. Enrichment at DNase I hypersensitive sites was observed across several implicated tissues and cell types (Fig. 3c), an observation that implicates diverse regulatory mechanisms with relevance across multiple tissue contexts.

Having identified loci associated with the latent factor, we next integrated orthogonal evidence at the variant, gene and phenotype-level annotations to nominate candidate genes (Methods). Using Polygenic Priority Score (PoPs) [39], we retained the highest ranked gene within each locus prioritizing 151 genes (Table S9). Colocalization [40] between the latent factor GWAS and expression quantitative trait locus (eQTL) catalogue [41] from 11 GTEx v8 [42] tissues relevant to both disease domains yielded 408 colocalizations across 151 unique genes (Table S10). Fine-mapping using PolyFun [43] coupled with SuSiE [44] (Table S11) and FINEMAP [45] (Table S12) identified 107 consensus variants at a posterior inclusion probability threshold (PIP)>0.5 mapped to genes through genomic proximity. Mouse knockout phenotypes obtained from the Mouse Genome Informatics (MGI) database [46] prioritized 276 genes involved in immune and cardiovascular-related phenotypes (Table S13), while human loss-of-function (LoF) evidence from the European Variation Archive (EVA) [47] identified candidates with direct evidence of altered gene function in humans (Table S14). This evidence was combined in an unweighted fashion to yield a final list of 473 genes identified by at least one evidence source (Table S15).

Genes underlying the latent genetic architecture may represent candidate pharmacological entry points in the psoriasis-CVD multimorbidity [48]. Combining discrete sources [49, 50] and excluding for psoriasis or cardiovascular adverse effects [51], we retrieved 663 drug-gene interactions (Table S16). Filtering for approved pharmacotherapies in both disease domains using the Open Targets Platform [52] (Table S17), we retained 90 approved compounds interacting with 23 candidate genes through 125 unique drug-gene relationships. Most drug-gene interactions were restricted to cardiovascular pharmacology including the LDL-targeting cluster (alirocumab, inclisiran, mipomersen, berberine and colestipol hydrochloride), the guanylate cyclase subunits *GUCY1A1*/*GUCY1A2*/*GUCY1B1* (riociguat, vericiguat, isosorbide mononitrate, nitric oxide and sodium nitroprusside), and the adrenergic receptor genes *ADRB1* and *ADRA1D* associated with approved HT and HF agents. Evidence for psoriasis-specific drug-gene interactions was restricted to *HFE* interacting with adalimumab and *TYK2* with deucravacitinib (Fig. 4). Eight genes interacted with compounds annotated for psoriasis and cardiovascular indications including six previously known *DOCK6*, *GDF15*, *GLP1R*, *HRH1*, *PRDM16* and *KEAP1* genes, as well as two genes not achieving genome-wide significance in individual GWASs, *DRD2* and *PDE4B*. We encourage readers to use our full results presented at Table S17 for drug-compound interactions of interest.

**Fig. 4.**
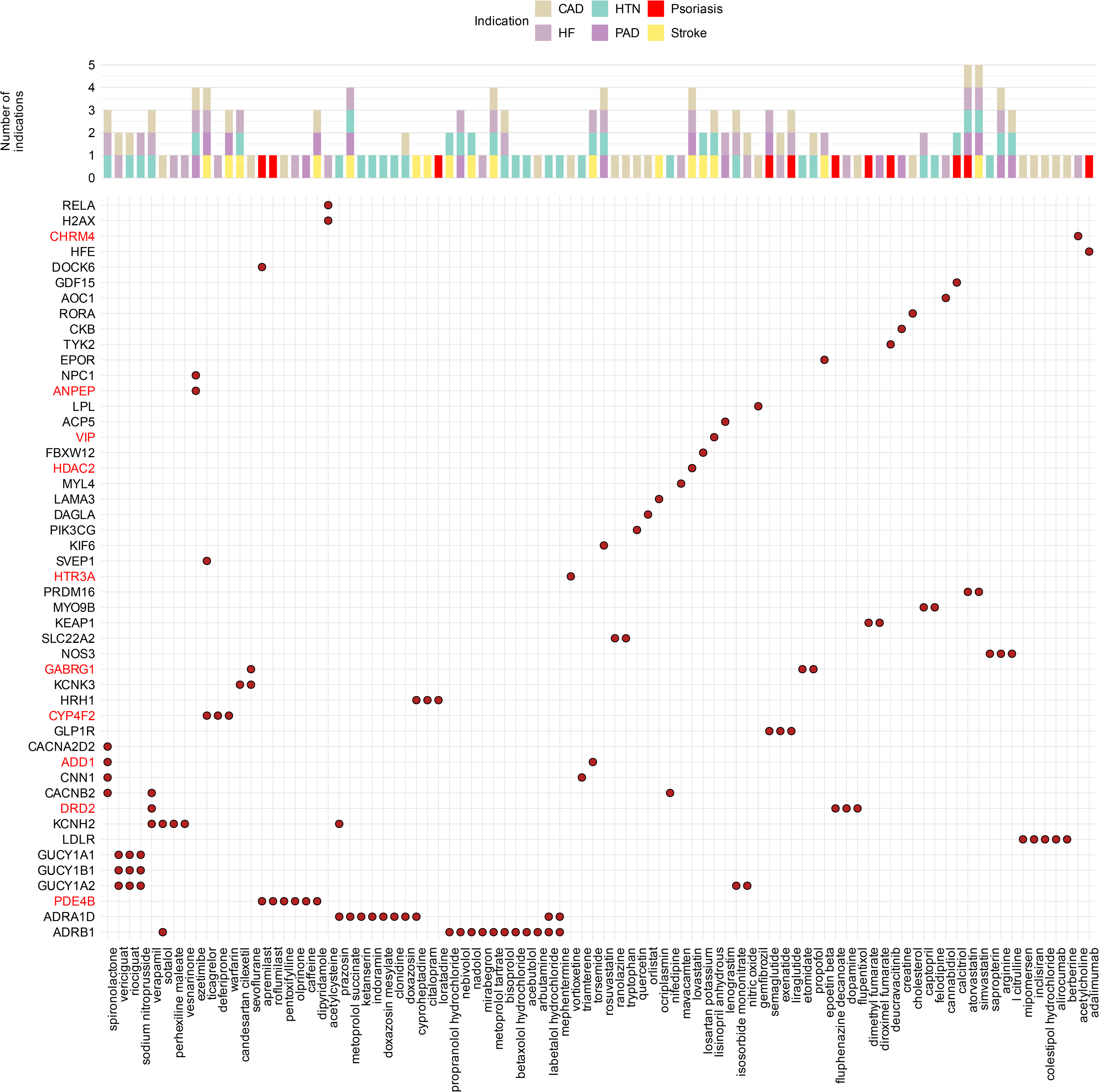
Druggable gene targets shared across psoriasis and cardiovascular disease risk loci. Dot plot showing known and investigational drugs (x-axis) mapped to their interacting genes (y-axis) at genomic risk loci associated with the latent psoriasis-cardiovascular disease (CVD) factor. Drug-gene pairs were curated by intersecting drug-gene interactions identified through the drug repurposing hub (DRH) and drug-gene interaction database (DGIdb) with Open Targets drug and disease-indication annotations, restricted to drugs with a recorded indication for hypertension (HT), heart failure (HF), psoriasis, coronary artery disease (CAD), peripheral artery disease (PAD), or stroke. Filled points indicate a drug-gene relationship. Genes and drugs are ordered by hierarchical clustering of shared drug-gene incidence. Gene labels shown in red correspond to genes at genomic risk loci identified from the latent psoriasis-CVD factor GWAS that did not show genome-wide significance in a ±250kb window around single-trait lead variants. The top panel summarizes the number of disease indications recorded across the CVD traits and psoriasis, colored by indication.

### Mediating pathways connecting psoriasis and cardiovascular diseases

To investigate mechanisms underlying shared genetic architecture, we considered five classes of putative mediating risk factors encompassing inflammatory markers [22,23], lifestyle risk factors [24,25,26,27,28], lipid traits [29], blood pressure [30] and metabolic markers [31,32,33,34,35], prioritizing well-powered GWASs of European ancestry (Table S1). Psoriasis showed nominal genetic correlation (P-value<0.05) with 17 of these traits. Mediation was quantified as attenuation of the standardized genetic association after conditioning on each individual mediator.

In the CVD-to-psoriasis direction, body mass index (BMI, 28.45%), waist-hip ratio (WHR, 23.94%) and CRP (28.59%) emerged as the strongest mediators, while lifestyle factors contributed only marginally with alcohol consumption and sleep duration each attenuating less than 2% of the association. In the psoriasis-to-CVD direction, type 2 diabetes (T2D) emerged as the stronger mediator (44.94%) (Fig. 5a). In both cases, even the strongest mediator explained less than half of the genetic association between psoriasis and CVD.

**Fig. 5.**
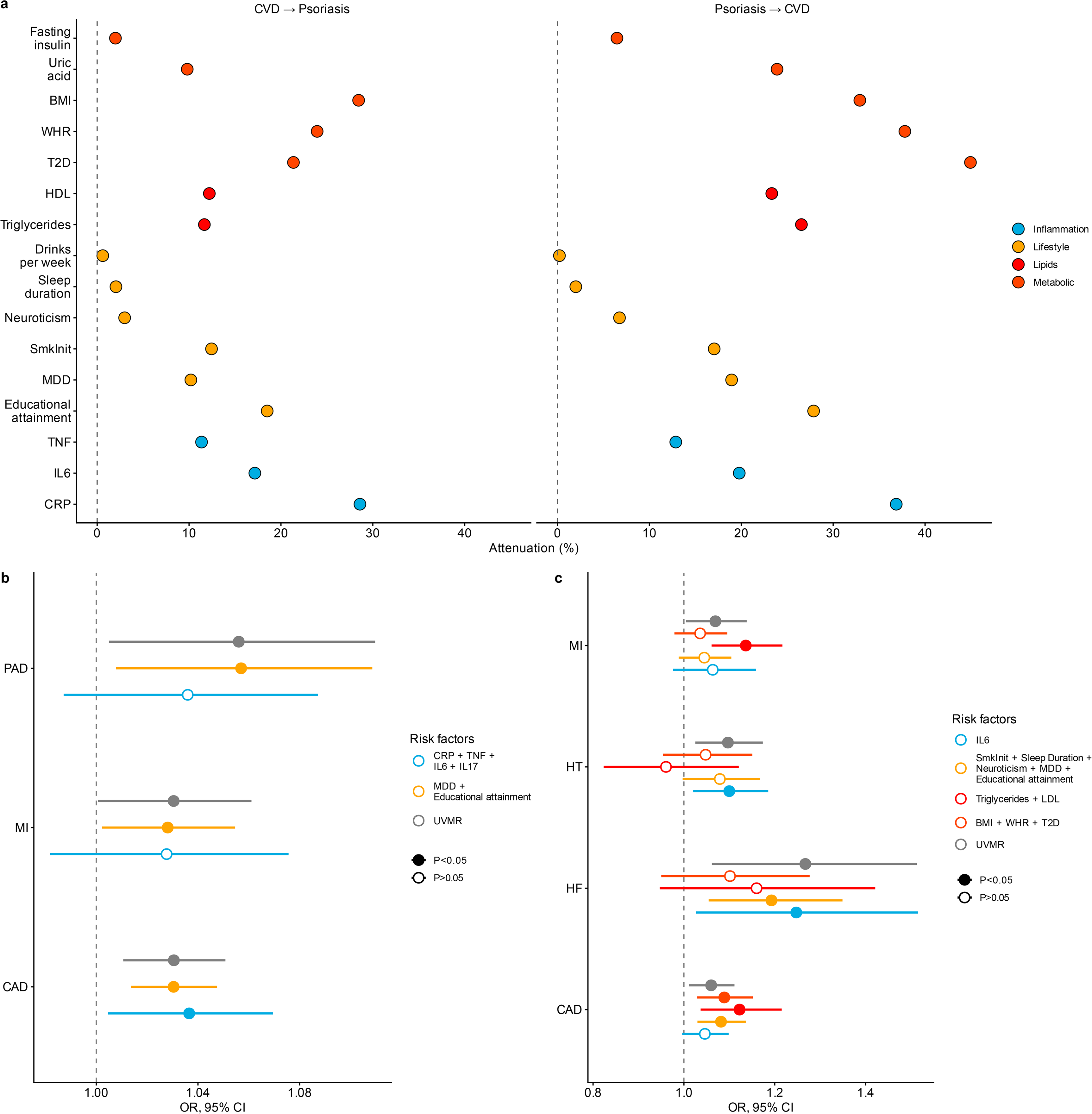
Shared risk factors and mediation analysis of the psoriasis-cardiovascular disease relationship. (a) Attenuation of the genetic association between psoriasis and cardiovascular disease (CVD) traits after conditioning on candidate mediators reporting nominal genetic correlation (P-value<0.05) with psoriasis. Points show the percentage attenuation of the psoriasis-to-CVD (left facet) and CVD-to-psoriasis (right facet) genetic association after adjusting for each candidate trait, colored by risk-factor category. The dashed vertical line indicates no attenuation (0%). (b, c) Multivariable Mendelian randomization (MVMR) estimates of the causal effect of (b) psoriasis on CVD outcomes and (c) candidate risk factors on psoriasis after jointly modeling shared risk factor pathways. Points show odds ratios (OR) with 95% confidence intervals from univariable MR (UVMR) and multivariable models adjusting for grouped risk factors (colored by risk factor grouping). Fill indicates whether the estimate reached nominal statistical significance (P-value<0.05). The dashed vertical line indicates the null (OR=1).

Attenuation estimates varied across CVD components (Table S18). In the psoriasis-to-CVD direction, HT showed the largest attenuation including T2D (77.8%), BMI (63.0%), WHR (62.2%) and CRP (60.6%). In the CVD-to-psoriasis direction, T2D (73.8%) and BMI (59.4%) showed the largest reported attenuation ratio estimates.

The latent psoriasis-CVD factor was positively correlated with blood pressure traits, adiposity measures and T2D (Table S7). Correlations were significant for triglycerides (r_g_=0.416) and HDL (r_g_=-0.408), but weak or non-significant correlations with LDL (r_g_=0.134, P-value=0.63) and total cholesterol (r_g_=0.097, P-value=2.37×10^-7^). Lifestyle risk factors contributed meaningfully, as is the case with major depressive disorder (MDD, r_g_=0.337, P-value=4.85×10^-71^). Inflammatory markers were positively correlated, though with considerably wider confidence intervals for the cytokine measures reflecting their comparatively underpowered GWASs.

Since attenuation is not proof of causal mediation, we complemented our analyses with bidirectional univariable MR (UVMR) and multivariable MR (MVMR) between psoriasis and each CVD component. Associations reaching nominal significance (P-value<0.05) in UVMR were submitted to MVMR analyses. Genetic liability to psoriasis was associated with CAD (odds ratio (OR), 95% CI, 1.03, 1.01-1.05), HF (OR, 95% CI, 1.025, 1.01-1.041), PAD (OR, 95% CI, 1.056, 1.005-1.109) and MI (OR, 95% CI, 1.03, 1.0007-1.061), while associations with AF, stroke and HT were null using the inverse-variance weighted (IVW) estimates (Table S19). Genetic liability to psoriasis also showed effects on educational attainment (OR, 95% CI, 0.992, 0.989-0.998) and all four inflammatory markers examined CRP, TNF, IL6 and IL17. The MR-Egger intercept showed evidence for horizontal pleiotropy between psoriasis and HF (P-value=0.033) (Table S20). For all other associations, effect directionality was consistent across MR-Egger, weighted median (Table S19) and MR-PRESSO (Table S21). In the CVD-to-psoriasis direction, IVW estimates associated genetic liability to CAD (OR, 95% CI: 1.06, 1.011-1.111), HF (OR, 95% CI: 1.267, 1.062-1.513), HT (OR, 95% CI: 1.097, 1.025-1.174) and MI (OR, 95% CI: 1.069, 1.005-1.138) with psoriasis. Among risk factors, we found effects on psoriasis risk from major metabolic traits (T2D, BMI, WHR), most lifestyle traits (smoking initiation, sleep duration, neuroticism, MDD and educational attainment), lipid traits (triglycerides, LDL) and IL6 (Table S19), without evidence for horizontal pleiotropy (Table S20) surviving outlier correction (Table S21).

In MVMR, the psoriasis-CAD estimate remained significant after separate adjustment for inflammatory markers and lifestyle factors. Estimates for MI and PAD were attenuated after adjusting for inflammatory markers (Fig. 5b). Educational attainment remained associated with all CVD outcomes, whereas CRP retained a significant independent effect only on PAD (Table S22). In the CVD-to-psoriasis direction, no single adjustment set explained all CVD-to-psoriasis associations (Fig. 5c). Metabolic adjustment attenuated the effects of MI and HT on psoriasis. Adjustment for IL6 attenuated the associations of CAD and MI with psoriasis, whereas lifestyle factors attenuated associations for HT and MI. Lipid adjustment attenuated the associations of HF and HT with psoriasis. Across models, BMI, educational attainment, smoking initiation and IL6 showed the most consistent independent associations while WHR, MDD, neuroticism and LDL effects in psoriasis were consistently attenuated to non-significance (Table S22). These findings suggest that adiposity and smoking behaviour may act as independent drivers of the bidirectional psoriasis-CVD relationship, while HF and CAD appear to be the CVD components with the strongest evidence of an independent association with psoriasis risk beyond shared metabolic, inflammatory and behavioural confounding (Fig. 5c).

## Discussion

In this study, we show that psoriasis-CVD multimorbidity is unlikely to be explained by a single inflammatory axis or conventional cardiovascular risk factors alone. Instead, psoriasis shares a functionally heterogeneous genetic architecture with multiple cardiovascular outcomes. This distinction is important because genome-wide correlations average effects across loci and may miss architectures in which many variants are shared but differ in magnitude, tissue context or direction of effect. Our findings therefore position the observed multimorbidity as a highly polygenic state rather than the consequence of a small number of pleiotropic loci.

The strongest regional signal at SH2B3-ATXN2 points to 12q24 as a pleiotropic region linking autoimmunity with cardiovascular outcomes as suggested elsewhere [53]. By contrast, the MHC showed only limited and mildly negative local correlations. Hence, cardiovascular multimorbidity in psoriasis does not appear to arise mainly from the major psoriasis susceptibility region but instead involves distinct and potentially non-immune signals relevant to multimorbidity. Consistently, psoriasis showed relatively sparse polygenicity but high discoverability and a large fraction of psoriasis non-zero variants were shared with CVDs (Fig. 2). This asymmetry of the genetic overlap corroborates clinical observations that psoriasis is associated with excess cardiovascular events beyond traditional risk factors [4]. Our results enhance the temporal and severity-dependent mechanisms proposed by the psoriatic march [6] through germline variation, indicating that part of the association may reflect shared bidirectional liability with interactions between discrete disease modalities such as immune activation, vascular dysfunction and cardiometabolic stress.

Previous studies primarily identified pleiotropic loci shared between psoriasis and individual CVDs [11,12,13,54,55]. Our study extends these observations by modelling a latent factor that captures the genetic liability common across psoriasis and multiple CVDs. This multivariate approach yielded 50 risk loci not achieving genome-wide significance in any individual GWAS. The exclusion of AF and MI from the best-fitting factor is informative, since AF has strong electrophysiological and atrial-remodeling components while MI reflects an acute atherothrombotic event that partially overlaps with chronic CAD liability. The poorer model fit indicates that psoriasis-related cardiovascular multimorbidity is more closely aligned with chronic vascular processes than with all cardiovascular endpoints equally. Functionally, the latent factor was enriched for a multi-tissue regulatory architecture (Fig. 3c), fitting the biology of psoriasis as a systemic inflammatory disease [56] and atherosclerosis as a chronic inflammatory process involving multiple cell types across different biological contexts [57]. Cumulatively, our results stress the added value of modeling shared liability directly instead of treating comorbidity as a secondary annotation of individual GWASs.

The drug-gene interaction analyses translated these observations into testable hypotheses with pharmacological relevance. Unlike conventional repurposing approaches that rely on phenotypic similarity or existing disease annotations, our analysis prioritized targets emerging from shared human genetic architecture (Fig. 4). Our results propose several translational entry points but they should be interpreted as genetic prioritization rather than therapeutic validation. Most nominated targets reflect established cardiovascular pharmacology, including lipid-lowering, guanylate-cyclase and adrenergic pathways. Others align closer with the psoriasis-CVD interface. *PDE4B* is notable since PDE4 inhibition modulates intracellular cAMP signaling and inflammatory cytokine production and apremilast is already used in psoriasis [58,59]. *GLP1R* is also attractive connecting exenatide and liraglutide with CAD and HF, while semaglutide provided additional evidence in psoriasis along with growing interest in GLP-1 receptor agonist across cardiometabolic and inflammatory disease [60,61]. Other candidates with cross-disease annotations provided complementary pharmacological hypotheses. For instance, *GDF15* linked with calcitriol, an approved drug for mild-to-moderate psoriasis with potential benefits in HT [62]. *HRH1* interacted with loratadine, an antihistamine effective in reducing pruritus in psoriasis patients [63]. *PRDM16* linked atorvastatin with evidence across psoriasis, CAD, HF, HTN and PAD to simvastatin, a drug approved for CAD and stroke. *KEAP1* interacted with two fumarate esters, dimethyl fumarate approved for psoriasis [64], and diroximel fumarate in early-phase clinical research for PAD. Finally, *DRD2* was linked to fluphenazine decanoate previously investigated in psoriasis (NCT00356200) and independently to dopamine, verapamil and flupentixol across multiple cardiovascular outcomes, consistent with proposed roles of dopaminergic signaling in keratinocyte biology and vascular function. Our results suggest that the therapeutic relevance of shared genetic liability may extend across disease domains rather than a single common pathway. However, further functional and clinical evidence is required to establish their therapeutic benefit in patients with psoriasis-CVD multimorbidity.

The mediation and MR analyses provided complementary evidence the excess cardiovascular risk associated with psoriasis cannot be explained solely by conventional cardiometabolic pathways. While consistent in both directions, even the strongest single mediators attenuated less than half of the shared genetic signal (Fig. 5a). The lipid profile of the latent multimorbidity factor further supports this interpretation. Strong genetic correlations with triglycerides but weaker associations with LDL and total cholesterol (Table S7) are more consistent with a dysmetabolic lipid profile than with classical cholesterol-driven atherosclerosis. HT was a clear exception, showing the most extensive attenuation estimates by metabolic and inflammatory traits in both directions (Table S18). The bidirectional associations derived from MVMR further argue against unidirectional approaches, suggesting that vascular and cutaneous pathogenic processes are reciprocally reinforced instead of one uniformly preceding the other. These results align with cardiovascular trials showing that inflammatory inhibition can reduce vascular events independently of lipid lowering [65].

Several limitations warrant consideration. First, our analyses were restricted to individuals of European ancestry to maximize statistical power, so generalizability to other ancestries remains unknown. Second, GWAS summary statistics reflect prevalent, lifetime genetic liability rather than dynamic relationships over disease course, limiting our ability to capture temporal associations. Third, despite removal of overlapping UK Biobank samples from the psoriasis GWAS (Methods), residual sample overlap with the mediating trait and CVD GWASs cannot be entirely excluded. However, previous studies have shown that bias introduced from sample overlap is minimal in large biobank-based settings [66]. Fourth, compared to the CRP GWAS, cytokine summary statistics remain comparatively underpowered which may underestimate mediation estimates. Finally, drug-target annotation identifies biological plausibility and repurposing opportunities but does not establish efficacy, safety or direction of therapeutic effect in patients with psoriasis-CVD multimorbidity.

Overall, our findings reframe the psoriasis-CVD multimorbidity arising from a largely concordant polygenic liability. This liability is neither fully explained by traditional risk factors nor is reducible to skin or vascular-derived biology alone. Our results connect epidemiological comorbidity with molecular mechanisms and highlight both known and novel candidates as starting points for mechanistic and therapeutic investigation that could benefit patients across the psoriasis-cardiovascular disease spectrum.

## Methods

### Data sources

We used publicly available GWAS summary statistics of European ancestry to maximize statistical power. All GWASs were retrieved from the GWAS catalog or resources provided by the respective publications. Details of all GWAS datasets, including sample size, ancestry, phenotype definition and source are provided in Table S1. For each retrieved GWAS, we harmonized variant identifiers, genomic coordinates, effect alleles and minor allele frequency (MAF) using the haplotype reference consortium (HRC) panel [67], genome build GRCh37/hg19. When necessary, genomic coordinates were transformed using the UCSC liftOver tool. To minimize sample overlap in genetic architecture and SEM analyses, we removed UKB participants from the psoriasis summary statistics using the MetaSubstract v1.6 R package [68]. The 1000 Genomes (1KGP) Europeans dataset was used as an LD reference panel [69].

Summary statistics for *cis*-eQTLs were retrieved from the eQTL catalogue [41]. We retrieved 11 tissues related to both psoriasis and CVDs, including adipose subcutaneous and visceral, artery aorta, coronary and tibial, Eppstein-Barr transformed lymphocytes, heart appendage and ventricle, sun exposed and not-sun exposed skin and whole blood from the GTEx v8 consortium [42]. All *cis*-eQTL datasets were transformed to GRCh37/hg19 coordinates using the UCSC liftOver tool.

All analyses were conducted using summary-level data with ethical approval and informed consent provided in the original studies.

### Shared polygenic architecture

#### Global and local genetic correlation

We estimated SNP-based heritability and pairwise genome-wide genetic correlations using LDSC [70]. Variants used as input to the LDSC were filtered for HapMap3 well-imputed variants in all cases. Pre-computed European LD scores from the 1KGP reference panel were used. Statistical significance for psoriasis-CVD genetic correlations was assessed using a Bonferroni threshold of P-value<0.05/7.

To determine whether global genetic correlations were driven by specific genomic regions, we estimated local genetic correlations using LAVA [36]. The genome was partitioned into 2,495 approximately independent LD blocks. For each psoriasis-CVD pair, local bivariate genetic correlations were estimated within each block using European ancestry LD information. Regions with insufficient local heritability or unstable covariance estimates were excluded according to LAVA default setting. An FDR-adjusted P-value<0.05 was considered significant.

### Extent of shared variants

We next employed MiXeR v1.3 to evaluate univariate polygenicity and bivariate genetic overlap between psoriasis and each CVD. MiXeR models GWAS summary statistics as a mixture of null and non-null genetic effects, estimating the number of variants with non-zero effects, the variance of effect sizes among non-zero variants, and the number of shared and trait-specific causal variants [37]. Univariate and bivariate models were fitted for psoriasis, CVD and psoriasis-CVD pairs. Associated variants were considered to explain at least 90% of the heritability estimated from common SNPs outside the MHC region, further referred to as causal variants. To increase robustness of point estimates, each pairwise comparison was fitted 20 different times for 2 million randomly selected variants with MAF≥0.05 at an r^2^ threshold of 0.8.

Model fit was assessed comparing the difference between the Akaike information criterion (AIC) for the best-fit MiXeR estimates and a reference model representing the minimum and maximum plausible degrees of polygenic overlap. A positive value of this difference validates the existence of the polygenic overlap beyond the minimal level required to explain the observed correlation estimates. The consistency of parameter estimates across the 20 independent runs was also examined to ensure model stability.

### Biological mechanisms

#### Construction of a multimorbidity factor

To appropriately model a genetic factor that represents the genetic liability between psoriasis and CVDs, we used genomic SEM v0.0.5 [16]. SNP covariance matrices and sampling covariance matrices were estimated from GWAS summary statistics using LDSC as implemented in genomic SEM, accounting for sample overlap. This genetic covariance matrix was next used to identify multivariate relationships. Criteria for acceptable model fit were set at CFI>0.95 and SRMR<0.05.

First, we modelled a latent factor that represents the genetic liability between implemented CVDs. The best model fit was constructed for a factor with CAD, HF, HT, PAD and stroke. For model identification purposes, the standardized loading of HF was set at 1. Next, we constructed a second-order latent factor that represents the genetic liability between CVDs and psoriasis, termed as psoriasis-CVD with psoriasis as indicator. The residual variance of the latent CVD factor was set to 0 to force all variance in the second-order latent psoriasis-CVD factor.

We conducted a multivariate GWAS on the psoriasis-CVD factor using the default diagonally weighted least squares estimator. To identify variants with heterogeneous effects across traits, we compared two alternative SEM specifications for each SNP, including a common pathway model and an independent pathway model, in which direct effects on each phenotype were estimated separately. We compared the common pathway (psoriasis-CVD) to the independent pathway to derive the heterogeneity measure Q_SNP_. For downstream analyses, SNPs with significant evidence of heterogeneity (Q_SNP_<0.05) were excluded. The effective sample size was calculated as 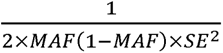, restricted to SNPs with 0.1≤MAF≤0.4, and estimated at 1642846. LDSC attenuation ratio was used to evaluate the extent to which inflation reflected polygenicity rather than confounding [71].

### Functional annotation and gene prioritization

To functionally annotate the multimorbidity psoriasis-CVD factor, we first identified independent genomic risk loci through FUMA using a two-stage clumping procedure [72]. Independent risk variants were first defined at r^2^<0.6 around a ±250kb window. Lead variants were then identified by clumping independent variants at an r^2^<0.1. This procedure was repeated for all individual GWASs used as input in the genomic SEM model. To identify loci not previously detected in included traits, the second-order latent factor loci were compared against genome-wide significant loci of all individual GWASs around a ±250kb window of each lead variant. Tissue enrichment analyses were performed across multiple P-value thresholds (P-value≤5×10^-8^, 10^-7^, 10^-6^, 10^-5^, 10^-4^ and 1) using GARFIELD v2 with default settings [38].

To prioritize candidate genes at each locus, we investigated all genes around ±250kb each lead variant. We collated evidence from 5 discrete annotation categories.

First, we used PoPS v0.2 method [39], a similarity-based gene prioritization framework that leverages genome-wide association signals together with diverse biological annotations. PoPS integrates various features including gene expression profiles, biological pathways, and protein-protein interaction networks to identify functional features enriched among genes associated with the trait and assigns a priority score to every protein-coding gene independent of local association statistics. The latent factor GWAS summary statistics were converted to gene-level associations using MAGMA v1.10 [73] using GRCh37/hg19 gene annotations extended by 10 kb. These gene-based associations results were submitted to PoPs. For each associated locus, the gene with the highest PoPS score was considered the leading candidate.

Second, colocalization analyses were performed to identify genes whose expression quantitative trait loci shared a likely causal variant with the latent psoriasis-CVD GWAS signal. For each gene-tissue pair within latent factor risk loci, we tested colocalization under a single causal variant assumption using coloc v5.2.3 [40]. Analyses used effect sizes and corresponding standard errors. Regional association summary statistics were evaluated under five competing hypotheses: no association (H0), association with only the first trait (H1), association with only the second trait (H2), association with both traits but distinct causal variants (H3), or association with both traits through a shared causal variant (H4). Gene-tissue pairs were considered colocalized when PP.H4/(PP.H3 + PP.H4)≥0.8.

Third, we performed functionally informed statistical fine-mapping for all latent factor risk loci using PolyFun coupled with SuSiE v0.14.2 [44] and FINEMAP v1.4.1 [45], following the Statistical and Functional Fine-mapping Applied to GWAS risk loci (SAFFARI) pipeline [74]. Functional prior causal probabilities were based on pre-computed baseline models derived from UK Biobank using 187 epigenomic and genomic annotations [75]. In the absence of an in-sample linkage disequilibrium (LD) reference panel, the UKB reference panel from the Alkes Group was used. Fine-mapping was performed independently with SuSiE and FINEMAP for each risk locus, defined as ±250kb around each lead variant. Variants with PIP>0.5 that belonged to a 95% credible set were retained and a consensus set was constructed across both methods. Fine-mapped variants were mapped to genes using genomic proximity.

Fourth, we incorporated orthogonal phenotype evidence from mouse knockout models. Mouse gene-phenotype associations were obtained from Mouse Genome Informatics (MGI) database for all genes mapped ±250 kb around each lead variant [46]. Prioritized genes were annotated for knockout phenotypes related to a dictionary constituted of cardiovascular traits and biology, skin and immune-related terms. Genes with relevant mouse phenotypes were retained as having model-organism support.

Fifth, human LoF evidence was obtained from the EVA [47]. Genes were prioritized if they harbored clinically annotated disruptive variants associated with relevant human phenotypes or disease annotations similarly to the MGI filtering procedure. This evidence was summarized in an unweighted fashion to prioritize genes at each locus.

Finally, we evaluated the therapeutic relevance of candidate genes using two complementary drug-target resources, the Drug Repurposing Hub (DRH) (8/19/2025 version) [49] and the Drug-Gene Interaction database (DGIdb) v5.0.11 [50]. Genes supported by at least one evidence layer were queried against both resources. In DGIdb, we retained gene-drug interactions with score>0.5. Drugs were retained only with regulatory approval or launched clinical status. Adverse drugs effects were identified using the above dictionary through the On-Label Side Effects (OnSIDES) v3.1.1 resource [51]. Therapeutic indications for all eligible drugs were retrieved from the Open Targets Platform [52]. Drug-gene pairs were harmonized by Ensembl gene ID and compound name and summarized by target gene, compound, indication and approval status.

### Modifiable risk factors linking psoriasis and CVDs

We used two complementary approaches to evaluate whether shared modifiable risk factors explain the association between psoriasis and CVDs. We first used our risk factors panel as mediators in the genomic SEM model. We required potential mediators to show nominal (P-value<0.05) genetic correlation with psoriasis. Analyses were performed in CVD-to-psoriasis and psoriasis-to-CVD directions. For each mediator, we calculated the proportion attenuated as the relative reduction in the standardized genetic association after adjustment. Mediation was assessed for the latent psoriasis-CVD relationship and for individual CVD components.

Since the mediation analysis relied on untestable assumptions, we employed bidirectional MR to evaluate putative causal relationships between psoriasis, individual CVD outcomes and candidate mediating traits. MR relies on three core assumptions, including (i) genetic instruments are robustly associated with the exposure, (ii) are not associated with confounders and (iii) influence the outcome only through the exposure [76]. Instrumental variables (IVs) were selected from genome-wide significant variants associated with each exposure at a P-value threshold of 5×10^-8^. For underpowered cytokine GWASs, including TNF, IL6 and IL17, we used a relaxed P-value threshold of 10^-5^. All variants were clumped using the 1KGP LD reference panel at an r^2^ threshold of 0.001. Palindromic variants with ambiguous allele frequencies were removed. Exposure and outcome summary statistics were harmonized to align effect alleles. In all cases, variants were Steiger filtered to ensure that IVs explained significantly more variance in the exposure than the outcome [77]. The primary univariable MR estimate was obtained using IVW [78]. Sensitivity analyses included MR-Egger, weighted median estimation and MR-PRESSO [79]. Horizontal pleiotropy was assessed using the MR-Egger intercept [80]. Associations reaching nominal significance (P-value<0.05) in the IVW estimate, concordance of effect sizes across sensitivity analyses and non-significant horizontal pleiotropy were carried forward to MVMR.

The MVMR framework was used to estimate the direct association of psoriasis with CVD outcomes after accounting for candidate risk factors [81]. In the psoriasis-to-CVD direction, psoriasis was modelled jointly with significant univariable mediators for each CVD outcome. In the CVD-to-psoriasis direction, psoriasis was modelled as the outcome, with each CVD exposure substituted into models adjusted for traits nominally associated with psoriasis in univariable MR. Separate models were fitted for each risk factor to avoid overfitting and collinearity. After separate IV selection for each exposure, variants were jointly clumped to retain independent SNPs available for every exposure. Outcome associations were extracted for the selected instruments and harmonized to ensure consistent effect allele orientation. Instrument directionality was evaluated using Steiger filtering. Multivariable direct effects were obtained using the IVW MVMR approach implemented in the MVMR v0.4 R package [81].

## Data availability

All GWASs used in the current study are publicly available from the respective publications provided at Table S1. Summary statistics for the latent psoriasis-CVD factor will be made publicly available after publication of the manuscript.

## Code availability

All software used in this study is freely and openly accessible. The LAVA R package can be found at https://github.com/josefin-werme/LAVA. The MiXeR v1.3 software can be found at https://github.com/precimed/mixer. The GenomicSEM v0.0.5 R package can be found at https://github.com/GenomicSEM/GenomicSEM. The SAFFARI fine-mapping pipeline can be found at https://github.com/mkoromina/SAFFARI. Mendelian randomization analyses relied on the publicly available R packages TwoSampleMR and MVMR R packages. Additional tools included FUMA (https://fuma.ctglab.nl/), Mouse genome informatics (https://www.informatics.jax.org/), EVA (https://www.ebi.ac.uk/eva/), Drug repurposing hub (https://repo-hub.broadinstitute.org/repurposing-app), Drug-gene interaction database (https://dgidb.org/) and OpenTargets (https://platform.opentargets.org/). All analysis scripts are available from the corresponding author upon reasonable request.

## Acknowledgments

This work uses data provided by patients and collected by the NHS as part of their care and support. The authors would like to thank all the participants of the cited genome-wide association studies and the respective authors for sharing their data.

## Funding sources

None declared.

## Author contributions

C .A. and Y.V. designed the research. C.A. conducted and visualized the results. C.A., E.G., I.T. and Y.V. interpreted the results. C.A. wrote the manuscript. E.G., I.T. and Y.V. critically revised the manuscript.

## Competing interests

The authors have no relevant financial or non-financial interests to disclose.

## Supplementary Tables

Table S1. Summary of GWASs used in the study. Table S2. Global genetic correlation estimates between all 32 traits included in the study. Table S3. Local genetic correlation estimates between psoriasis and CVDs. Table S4. Results for the univariate MiXeR model for psoriasis and CVDs. Table S5. Results for the bivariate MiXeR model between psoriasis and CVDs. Table S6. Independent genomic risk loci of the latent psoriasis-CVD factor derived from FUMA. Loci in yellow did not overlap in genome-wide significant loci defined as 500kb around lead variants of individual GWASs. Table S7. Global genetic correlation estimates between the latent psoriasis-CVD factor and all 32 traits included in the study. Table S8. Tissue enrichment analysis using GARFIELD for the latent psoriasis-CVD factor. Table S9. Gene prioritization results for the latent psoriasis-CVD factor using PoPs. Table S10. Gene prioritization results for the latent psoriasis-CVD factor using colocalization in GTEx v8 tissues. Table S11. Fine-mapping results using PolyFun + Susie for the latent psoriasis-CVD factor. Table S12. Fine-mapping results using PolyFun + FINEMAP for the latent psoriasis-CVD factor. Table S13. Gene prioritization results for the latent psoriasis-CVD factor using Mouse Genome Informatics database. Table S14. Gene prioritization results for the latent psoriasis-CVD factor using the European Variation Archive and related clinical phenotypes. Table S15. Gene prioritization scores for all genes across independent risk loci. Table S16. Gene-drug interactions retrieved from the DGIdb and DRH databases. Table S17. Drug indications for psoriasis and related CVDs retrieved from the OpenTargets database. Table S18. Mediation analysis between psoriasis, related CVDs and the latent CVD factor. Table S19. Results for all univariable Mendelian Randomization analyses. Table S20. Results for the MR-Egger intercept sensitivity analysis. Table S21. Results for the MR-PRESSO sensitivity analysis. Table S22. Multivariable Mendelian Randomization results.

## Notes

### Competing Interest Statement

The authors have declared no competing interest.

